# Effect of polymerized type I collagen in hyperinflammation of adult outpatients with symptomatic COVID-19: a double blind, randomised, placebo-controlled clinical trial

**DOI:** 10.1101/2021.05.12.21257133

**Authors:** Silvia Méndez-Flores, Ángel Priego-Ranero, Daniel Azamar-Llamas, Héctor Olvera-Prado, Kenia Ilian Rivas-Redondo, Eric Ochoa-Hein, Andric Perez-Ortiz, Estefano Rojas-Castañeda, Said Urbina-Terán, Luis Septién-Stute, Thierry Hernández-Gilsoul, Adrián Andrés Aguilar-Morgan, Dheni A. Fernández-Camargo, Elizabeth Olivares-Martínez, Diego F. Hernández-Ramírez, Gonzalo Torres-Villalobos, Janette Furuzawa-Carballeda

## Abstract

**BACKGROUND:** Currently, therapeutic options for ambulatory COVID-19 patients are limited.

**OBJECTIVE:** To evaluate the safety, efficacy and effect of the intramuscular administration of polymerized type I collagen (PTIC) on hyperinflammation, oxygen saturation and symptom improvement in adult outpatients with symptomatic COVID-19.

**DESIGN:** Double-blind, randomised, placebo-controlled clinical trial of PTIC vs placebo.

**SETTING:** Single Third-level hospital in Mexico City (Instituto Nacional de Ciencias Médicas y Nutrición Salvador Zubirán)

**PARTICIPANTS:** Eighty-nine adult participants with a confirmed COVID-19 diagnosis and symptom onset within the 7 days preceding recruitment were included from August 31, 2020 to November 7, 2020 and followed for 12 weeks. Final date of follow-up was February 4, 2021.

**INTERVENTIONS:** Patients were randomly assigned to receive either 1.5 ml of PTIC intramuscularly every 12 h for 3 days and then every 24 h for 4 days (n=45), or matching placebo (n=44).

**MAIN OUTCOMES AND MEASURES:** The primary outcome was a mean reduction of at least 50% in the level of IP-10 compared to baseline. The secondary outcomes were mean oxygen saturation ≥92% while breathing ambient air and duration of symptoms.

**RESULTS:** Of 89 patients who were randomised, 87 (97.8%) were included in an intention-to-treat analysis; 37 (41.6%) were male and mean age was 48.5±14.0 years. The IP-10 levels decreased 75% in the PTIC group and 40% in the placebo group vs baseline. The comparison between treatment vs placebo was also statistically significant (*P=*0.0047). The IL-8 (44%, *P=*0.045), M-CSF (25%, *P=*0.041) and IL-1Ra (36%, *P=*0.05) levels were also decreased in the PTIC group vs baseline. Mean oxygen saturation ≥92% was achieved by 40/44 (90%), 41/42 (98%) and 40/40 (100%) of participants that received PTIC at 8, 15 and 97 days of follow-up vs 29/43 (67%), 31/39 (80%) and 33/37 (89%) of patients treated with placebo (*P*=0.001). The unadjusted accelerated failure time model showed that patients treated with PTIC achieved the primary outcome 2.70-fold faster (*P*<0.0001) than placebo. In terms of risk, the group of patients treated with PTIC had a 63% lower risk of having a mean oxygen saturation <92% vs placebo (*P*<0.0001). Symptom duration in patients treated with PTIC was reduced by 6.1±3.2 days vs placebo. No differences in adverse effects were observed between the groups at 8, 15 and 97 days of follow-up.

**CONCLUSIONS:** In this study, treatment with PTIC down-regulated IP-10, IL-8, M-CSF and IL-Ra levels, which could explain the PTIC effect on the higher proportion of patients with mean oxygen saturation readings ≥92% and a shorter duration of symptoms as compared to patients treated with placebo. Although results are encouraging, larger randomised trials are needed.

**TRIAL REGISTRATION:** ClinicalTrials.gov Identifier: NCT04517162

## Introduction

Coronavirus disease 2019 (COVID-19) is caused by the severe acute respiratory syndrome coronavirus 2 (SARS-CoV-2). Most symptomatic people have only a mild disease, but approximately 10–15% of patients have a moderate or severe disease that requires admission to hospital, oxygen support and/or admission to an Intensive Care Unit (ICU).^1^ Host inflammation, leading to pulmonary oedema and a syndrome that resembles acute respiratory distress syndrome (ARDS), occurs in the most severe cases^2^ and is a result of an exuberant activation of mononuclear cells that is accompanied by an elevation of inflammatory cytokines (cytokine-release syndrome, CRS) and chemokines (IL-1β, IL-1Ra, IL-2, IL-2Rα, IL-6, IL-7, IL-8, IL-9, IL-10, IL-13, IL-15, IL-17, IL-18, G-CSF, M-CSF, IFN-α2, IFN-γ, TNF-α, TRAIL, basic FGF, HGF, PDGF-BB, VEGF, GRO-α, IP-10, MCP-1, and MIG), inefficient production of type 1 interferons and impaired antiviral response. Left untreated, multiorgan failure and death could occur in patients with severe disease.^2,3^

Although dexamethasone is approved for the treatment of CRS in hospitalized patients with COVID-19 because it reduces the risk of death,^4^ non-hospitalized patients do not benefit from this therapy. Nonetheless, considering that clinical deterioration typically occurs during the second week of illness, an opportunity for prevention or mitigation of severe cases with early therapy exists; however, effective treatment of outpatients is an unmet need so far.

A potential drug for treatment of COVID-19 patients is the polymerized type I collagen (PTIC). It is a γ-irradiated mixture of pepsinised porcine type I collagen and polyvinylpyrrolidone (PVP) in a citrate buffer solution. At 37°C and neutral pH, the molecule does not form a gel, like collagen does, and its electrophoretic, physicochemical and pharmacological properties are modified by the covalent bond between the protein and the PVP moiety.^5^

Previous works have demonstrated that PTIC has immunomodulatory properties. The addition of 1% PTIC to synovial tissue cultures from patients with rheumatoid arthritis or osteoarthritis downregulates the following: pro-inflammatory cytokines (IL-1β, TNF-α, IL-8, IL-17, IFN-γ, PDGF and TGF-β1); the expression of adhesion molecules such as endothelial leukocyte adhesion molecule (ELAM-1), vascular cell adhesion molecule (VCAM-1) and intercellular adhesion molecule (ICAM-1); the expression of cyclooxygenase (Cox)-1 enzyme; and the collagenolytic activity through the modulation of transcription of factor NF-kB. ^6-15^ Moreover, PTIC has been shown to induce a positive regulation over the tissue inhibitor of metalloproteases (TIMP-1), the production of IL-10, and the presence of regulatory T cells.^11,14^

Previous clinical experience has shown that the intramuscular or subcutaneous administration of PTIC to patients with active rheumatoid arthritis, as a therapeutic co-adjuvant of methotrexate (phase II studies), produced a statistically significant improvement in the count of swollen joints and morning stiffness; 57% of patients achieved an ACR score of 50, and 30% had disease remission with this therapeutic combination. PTIC was safe and well tolerated in long-term treatment, without adverse effects.^16-19^

The main objective of this study was to evaluate the efficacy and safety of the intramuscular administration of PTIC on the hyperinflammation, defined as CRS, oxygen saturation and symptom improvement in adult symptomatic outpatients with confirmed COVID-19.

## Methods

### Study design

This was a single centre, double-blind, placebo-controlled, randomised clinical trial that compared PTIC to placebo in adult outpatients with confirmed COVID-19. The study was approved by the institutional review board of the Instituto Nacional de Ciencias Médicas y Nutrición Salvador Zubirán (INCMNSZ, reference no. IRE 3412-20-21-1; supplement 1) and was conducted in compliance with the Declaration of Helsinki,^20^ the Good Clinical Practice guidelines, and local regulatory requirements. All participants provided written informed consent. This study is registered with the ClinicalTrials.gov identifier NCT04517162.

Trial candidates were identified in a prospective database of patients that went to a medical appointment at the hospital and were discharged home with a diagnosis of COVID-19 and symptomatic treatment. Diagnosis was based on suggestive symptoms (fever, headache, cough or dyspnoea, plus at least another symptom such as malaise, myalgias, arthralgias, rhinorrhoea, throat pain, conjunctivitis, vomiting or diarrhoea) and a positive real-time reverse-transcription polymerase chain reaction result for SARS-CoV-2.

Staff reached candidates via telephone calls and informed them about the purpose of the study. Once in the hospital study site, inclusion and exclusion criteria were verified. Subjects that fulfilled the criteria mentioned above were included. Exclusion criteria were: hypersensitivity to PTIC or any of its excipients; COVID-19 patients that required hospitalization; all pregnant or breast-feeding women; patients with chronic kidney disease (estimated glomerular filtration rate less than 60 for more than 3 months or need for haemodialysis or hemofiltration); decompensated liver cirrhosis; congestive heart failure (New York Heart Association class III or IV); and patients with cerebrovascular disease, autoimmune disease, cancer, multiorgan failure or immunocompromise (solid organ transplant recipient or donor, bone marrow transplant recipient, AIDS, or treatment with biologic agents or corticosteroids). Study subjects were recruited between August 31 and November 7, 2020. Patients signed the informed consent before being randomly allocated to either PTIC or matching placebo.

During the first day of enrolment, candidates received a study kit that consisted of PTIC or placebo, a pulse oximeter, and a symptom questionnaire booklet. Patients were instructed on how to use the study medication and the pulse oximeter, and how to complete the questionnaires. Also, staff administered the first dose of PTIC or placebo on site, then participants self-administered their assigned treatment on their own.

Phone calls were made daily during the first 3 days of the trial to address participants’ questions, address any medication-related issues, and encourage completion of questionnaires daily. Additional phone calls were conducted on a case-by-case basis when participant’s survey data indicated values outside of the expected ranges. For participants that had a worsening disease course (89% or lower oxygen saturation while breathing ambient air), study staff recommended that medical attention be provided in the Emergency Department at no cost. If the patient required hospitalization and treatment with dexamethasone, then he/she was eliminated from the study. However, these patients were included in the intention-to-treat analysis. Patients were evaluated by staff at the study site (S.M-F, A.P-R, D.A-Ll, H.O-P, E.O-H, E.R-C.) on days 8, 15 and 97 (1, 7 and 90 days after the last dose of PTIC or placebo, respectively), and patients were encouraged to complete questionnaires daily.

### Participants and data collection

The study included non-hospitalized adults with COVID-19 whose symptoms started within the previous 7 days before randomization (fig 1). Individuals were asked to provide personal information (date of birth, type of job, educational level, previous contact with infected individuals), pre-existing conditions (systemic hypertension, diabetes mellitus, cardiovascular disease, cerebrovascular disease, hypertriglyceridemia, dyslipidaemia) and symptoms. Personal data, exposure history, clinical presentation, chest computed (CT) tomography, laboratory tests, previous treatment and outcome data were collected both prospectively and from inpatient medical records. Laboratory data collected from each patient from study days 1 (baseline), 8 (day 1 post-treatment), 15 (day 8 post-treatment) and 97 (day 90 post-treatment) included complete blood count, coagulation profile, serum biochemical tests (including renal and liver function tests, electrolytes, lactate dehydrogenase, D dimer and creatine kinase), serum ferritin, C-reactive protein (CRP) and procalcitonin. Chest CT scans were done in all patients at baseline.

**Figure 1.**
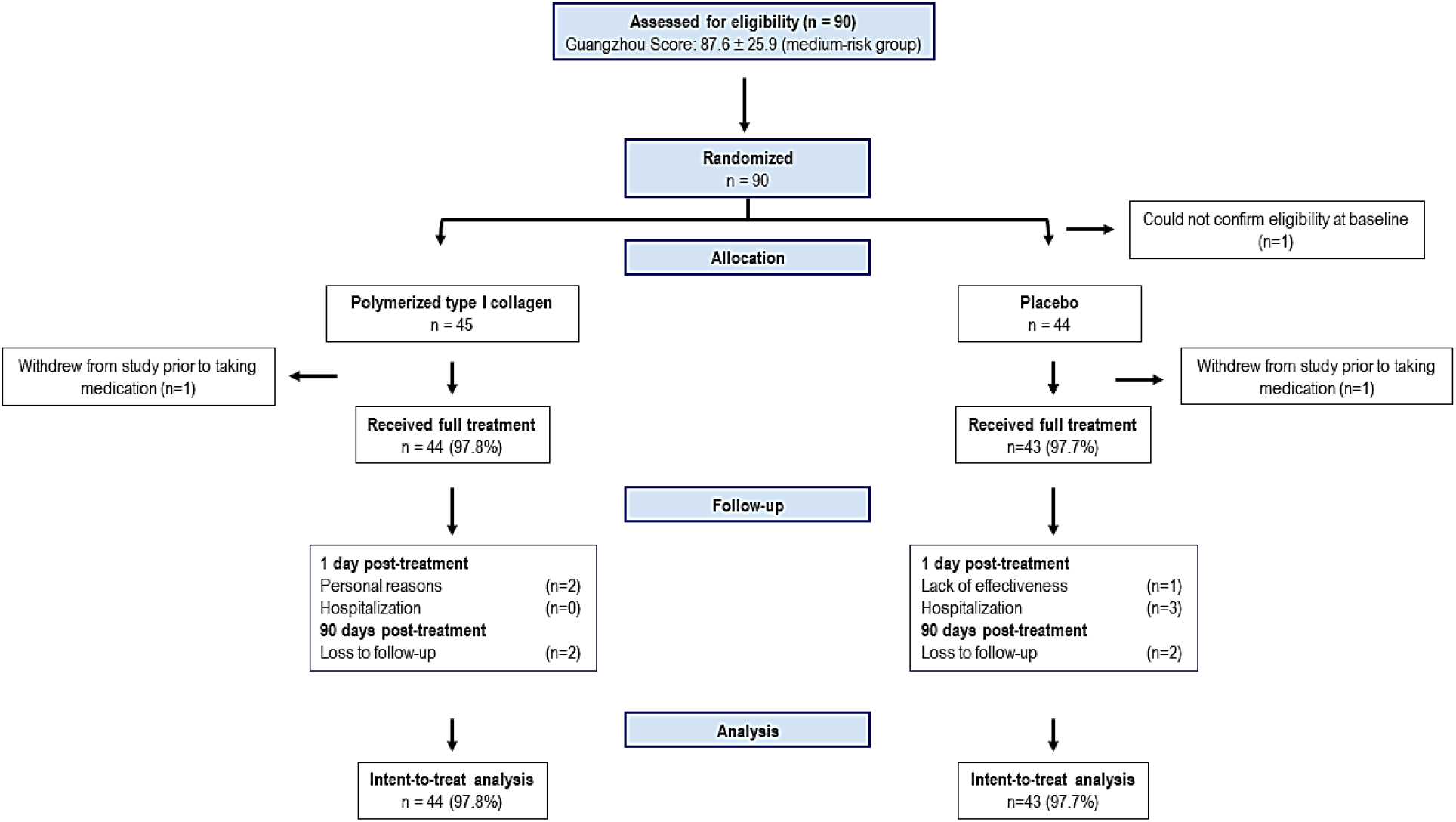
Flow chart.

### Study sample

According to a study published by Yang Y. et al,^21^ IFN-γ-inducible protein 10 (IP-10), a host protein involved in lung injury from virus-induced hyperinflammation, is highly associated with disease severity. IP-10 predicts the progression of COVID-19 and its reduction by 50% avoids the development of critical or severe disease. Thus, it was arbitrarily decided that a difference of 50% on this biomarker on day 8 (day 1 post-treatment) vs baseline would be significant. Assuming this difference as the effect size, two-tailed alpha=0.05, power=0.80 and Cohen’s d = 0.61522 and losses to follow-up, the sample size was set at 45 patients in each group. To calculate the sample size, the R software (version 3.6.2) with the “pwr” package was used.

### Randomization

Patients were randomised in a 1:1 fashion to PTIC or placebo. The Excel program displayed a random allocation list to the laboratory manager (K.R-R) who was in charge of assigning PTIC or placebo to study subjects. All outcome assessors, investigators and research staff who were in contact with participants were blinded to participant treatment assignment.

### Intervention

Participants received an intramuscular dose of either PTIC (1.5 ml, equivalent to 12.5 mg of collagen) every 12 h for 3 days and then every 24 h for 4 days, or placebo. Only acetaminophen or acetylsalicylic acid were allowed as concomitant therapy. Monitoring of compliance was evaluated by counting empty vials returned on subsequent visits.

### Measurement of serum cytokines

The blood samples were taken from study subjects at baseline and day 8 (1 day after the last dose of PTIC or placebo, respectively) and placed in serum separator tubes, followed by five tube inversions. The samples were allowed to stand for 20 minutes and were then centrifuged at 2000 rpm at 4°C. The sera were aspirated and separated into 300 μl aliquots. After collection, the serum samples were stored at -70°C. The concentrations of cytokines and chemokines were determined with the Hu Cyto screening panel 48-Plex kit (Bio-Rad, Berkeley, Calif) as reported elsewhere.^22^ According to their functions, the cytokines/chemokines were grouped into 6 categories: interleukins (IL-1α, IL-1β, IL-1Ra, IL-2, IL-2Rα, IL-3, IL-4, IL-5, IL-6, IL-7, IL-8, IL-9, IL-10, IL-12p40, IL-12p70, IL-13, IL-15, IL-16, IL-17, and IL-18), colony stimulating factors (G-CSF, GM-CSF, M-CSF, and stem cell factor [SCF]), interferons (IFN-α_2_ and IFN-γ), tumour necrosis factor (TNF-α, TNF-β, and TNF-related apoptosis inducing ligand [TRAIL]), growth factors (basic fibroblast growth factor [FGF], nerve growth factor-β [NGF-β], hepatocyte growth factor [HGF], leukaemia inhibitory factor [LIF], platelet-derived growth factor [PDGF-BB], vascular endothelial growth factor [VEGF], and stem cell growth factor-β [SCGF-β]), and chemokine family (cutaneous T-cell-attracting chemokine [CTACK], eotaxin, growth related oncogene-α [GRO-α], IFN-γ inducible protein-10 [IP-10], monocyte chemoattractant protein-1 [MCP-1], MCP-3, migration inhibitory factor [MIF], monokine induced by interferon-γ [MIG], macrophage-inflammatory protein [MIP-1α], MIP-1β, regulated upon activation normal T-cell expressed and secreted [RANTES], and stromal cell-derived factor-1α [SDF-1α]).

### Primary and secondary outcomes

The primary outcome was a mean reduction of at least 50% in the level of IP-10 compared to baseline. The secondary outcomes were an oxygen saturation of 92% or greater while breathing ambient air and duration of symptoms. Grading of symptoms was as follows: 0= absent, 1= mild, 2= moderate and 3= severe (continuous variable). Two comparisons were made regarding symptom duration and intensity: 1) at 8, 15 and 97 days as compared to baseline in each group, and 2) PTIC vs placebo.

The secondary outcomes were measured using participants’ self-reported responses during the next 8, 15 and 97 days after randomization. Each participant’s diary and health status were further checked by phone contact and by physician in-person evaluation on days 8, 15 and 97.

Adverse events and serious adverse events were intentionally sought by participants and study staff during the first 97 days after first dose of either PTIC or placebo (supplement 2, supplementary tables 1 and 2).

### Chest CT

A semi-quantitative scoring system was used to estimate the pulmonary involvement based on the pulmonary affected area.^23,24^

### Severity of the Guangzhou score

The occurrence of critical illness was determined according to the Guangzhou score.^25^

### Statistical analysis

A descriptive analysis was done: continuous variables were expressed by means and standard deviations (normal distribution) or medians and interquartile ranges (non-normal distribution), and categorical variables were summarized using proportions. The comparison of continuous variables between two groups was performed using the Wilcoxon rank sum test. For evaluation of the primary and secondary outcomes, intention-to-treat inferential analyses were done. Variables with a significant deviation from normality (set at the 0.05 level) in the Kolmogorov – Smirnov test were transformed to a logarithmic scale. In this comparison, the average differences represented the percentage of change (delta) between post-treatment status vs pre-treatment status in each study group. For continuous variables with normal distribution, delta was calculated by subtracting the pre-treatment value from the post-treatment value (on days 8, 15 or 97) and dividing by the pre-treatment value. Either Student’s t test, Wilcoxon rank sum test or Chi-squared (Pearson’s) test were used for the intention-to-treat inferential analysis of normal or transformed (normalized) continuous variables, non-normally distributed continuous variables, and categorical variables, respectively. Based on the post-intervention evaluation intervals (on days 8, 15 or 97), we implemented an exponential model of Accelerated Time Failure in SAS v 9.4. We adopted this approach based on the shape and scale parameters of the latter (δ ≈ σ ≈ 1, respectively). In addition, we analysed the other risk distributions and further corroborated the goodness of fit of the exponential by means of likelihood ratio tests, the Akaike information criterion (AIC), and the examination of logarithmic survival plots. For the Kaplan-Meier curves, we modelled oxygen saturation ≤92%-free survival in the PRISM GraphPad Statistics analysis program. To bolster our approach, we estimated the survivor function for the probability of achieving ≥92% oxygen saturation while breathing ambient air by the Kaplan-Meier method. Significant differences in survivor experience across the follow-up were corroborated with a log-rank test. Finally, we estimated the hazard for not meeting the primary outcome by Cox regression. Our data and model hold the proportionality of the hazard’s assumption.

## Results

### Baseline description of the study population

Eighty-nine adult non-hospitalized patients with COVID-19 (mild to moderate disease) were enrolled in the study (eligibility of one patient at baseline could not be confirmed and this patient was therefore excluded). The mean (±SD) age of the patients was 48.5±14.0 years. Thirty-seven patients (41.6%) were male. According to the Guangzhou score to predict the occurrence of critical illness, the mean score was 87.6±25.4 (medium risk). The mean (±SD) oxygen saturation of study participants was 92.5±2.5. Twenty-nine patients (32.6%) had oxygen saturation of 91% or lower while breathing ambient air (13 of the PTIC group and 16 of the placebo group). Fifty-one patients (57.3%) had a history of exposure to suspected or confirmed COVID-19 patients, 37 (41.6%) had sudden symptom onset, 33 (37%) had dyspnoea, and 28 (31.5%) had been vaccinated against influenza. Coexisting conditions and symptoms are described in table 1.

**Table1.**
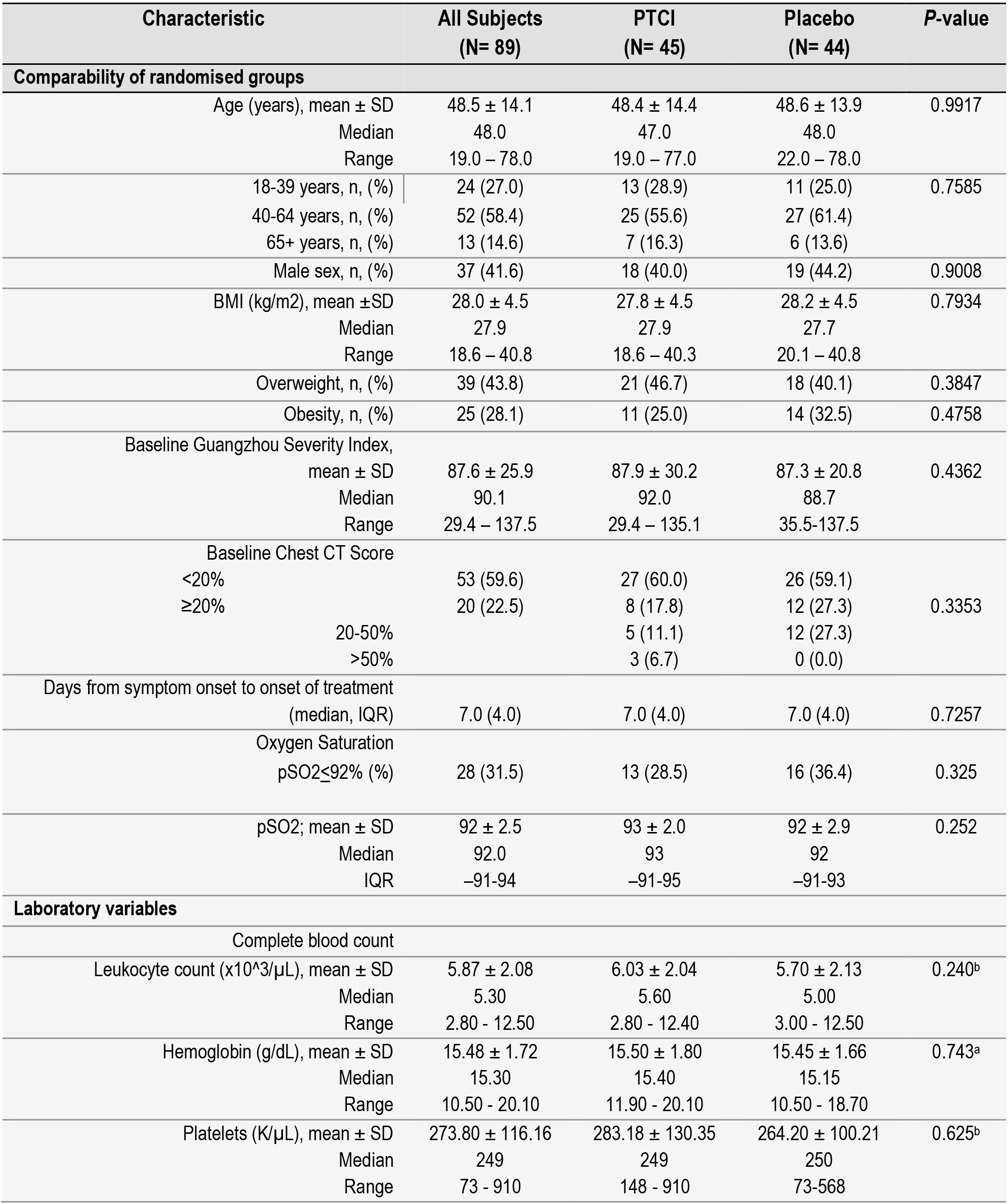
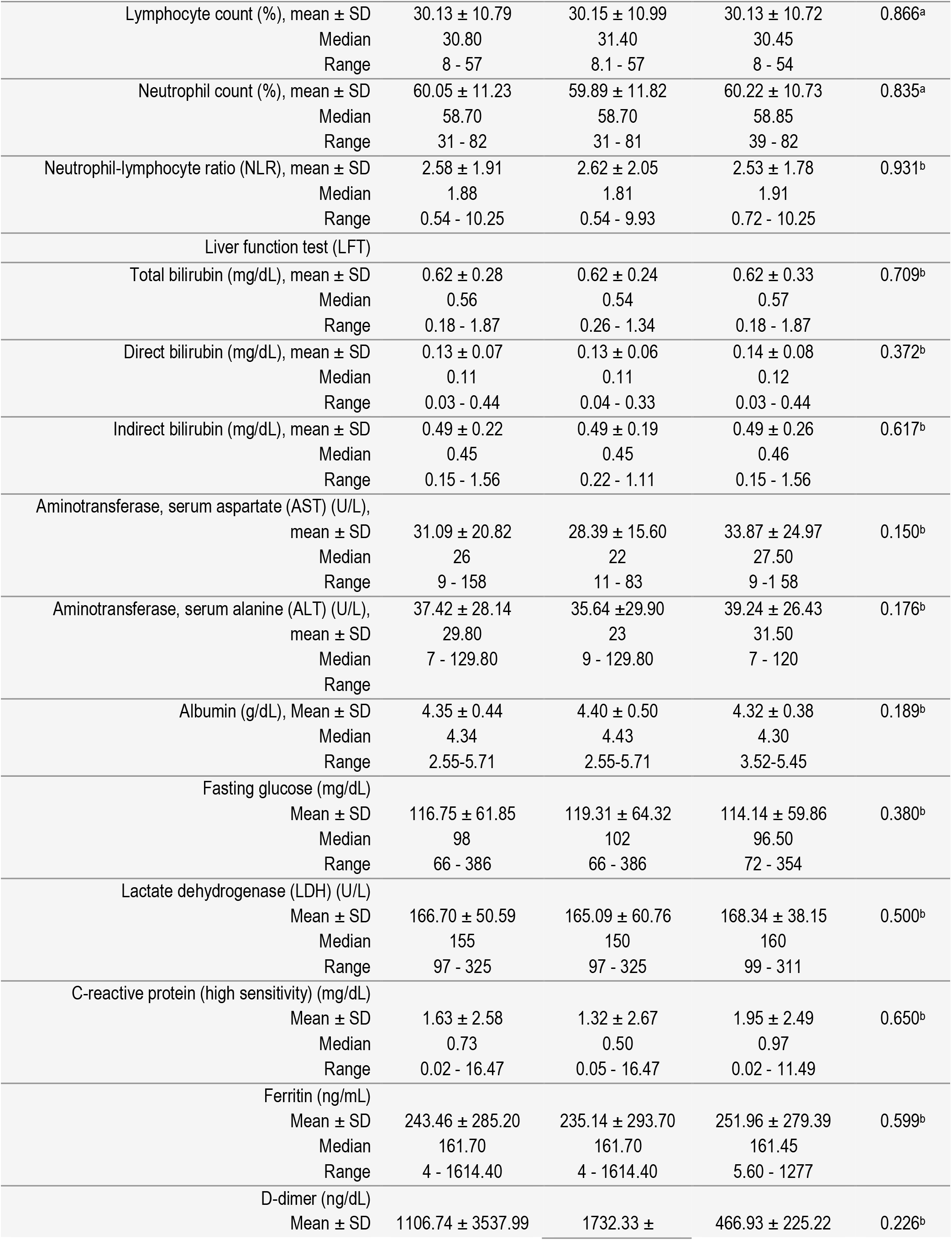
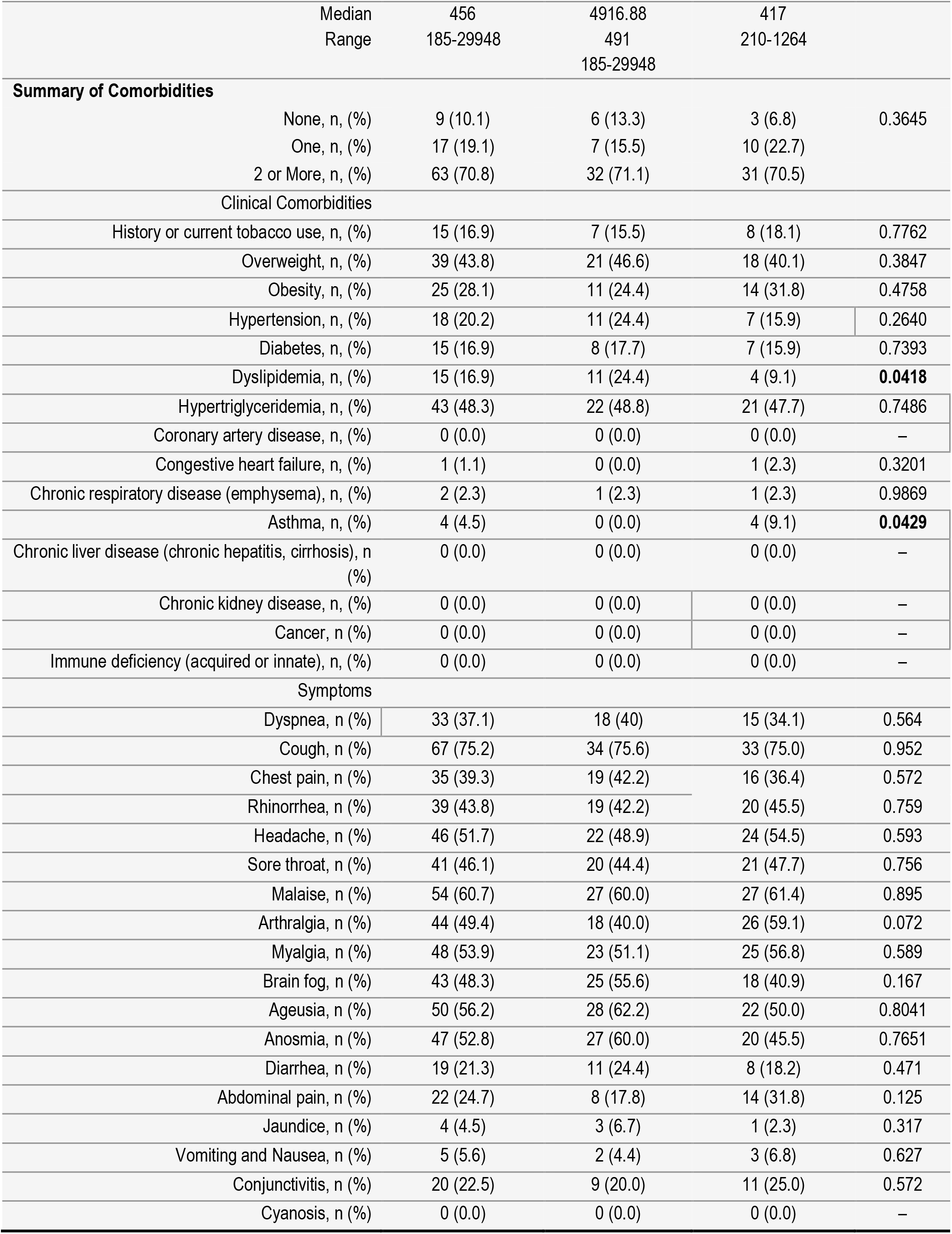
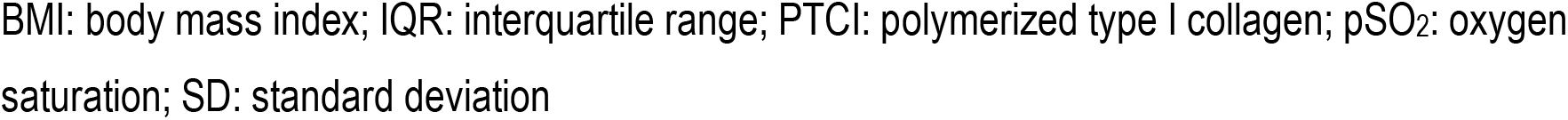
Baseline demographic and clinical characteristics of the trial population.

Regarding radiological abnormalities on chest CT, 73 patients (82%) had lung disease; of these, 53 (60%) had less than 20% lung parenchymal involvement, 17 (19%) had between 20 and 50%, and 3 (3%) had higher than 50% (Table 1).

Ninety-eight percent of patients in the PTIC group and 95.5% in the placebo group were analysed by the intention-to-treat principle. During week 1 of the study, one patient in the PTIC group withdrew from the protocol due to personal reasons (a psychiatric patient suffering from anxiety attacks), while two patients in the placebo group were withdrawn (eligibility could not be confirmed for one, and lack of effectiveness was present in the other). During week 2 of the study, 2 patients in the PTIC group withdrew from the protocol for personal reasons, while 3 patients in the placebo group were hospitalized (patients were eliminated from the study when they began treatment with dexamethasone). However, these patients were included in the intention-to-treat analysis. At 97 days, 2 patients in each group were lost to follow-up (fig 1).

### Concomitant medications

Of 89 patients at baseline, 64 (72%) were being treated with acetaminophen, 28 (31.5%) with acetylsalicylic acid, 5 (5.6%) with antivirals (oseltamivir, rimantadine), and 36 (40.4%) with antibiotics (azithromycin, ceftriaxone, penicillin, clarithromycin and levofloxacin). The use of acetaminophen (71% *vs*. 73%), acetylsalicylic acid (27% *vs*. 39%), antivirals (7% *vs*. 5%) and antibiotics (40% *vs*. 41%) were similar in the PTIC and placebo groups, respectively. No patients were treated with anticoagulants or steroids.

### Primary outcome

At day 8 post-treatment, the IP-10 levels decreased 75% in the PTIC group (*P<*0.001) and 40% in the placebo group (*P=*0.015) vs baseline; this reduction was greater in the former group than in the latter (*P=*0.0047; fig 2A). The IL-8 (44%, *P=*0.045), M-CSF (25%, *P=*0.041) and IL-1Ra (36%, *P=*0.05) levels were also decreased in PTIC group vs baseline (fig 2B-D). TRAIL levels were decreased in the placebo group (14%, *P=*0.002) vs baseline (fig 2E). IL-1α, IL-2, IL-5, IL-6, IL-10, IL-12p40, IL-12p70, IL-15, GM-CSF, VEGF, bNGF, IFN-α2, and MCP-3 were not detected at baseline or at day 8 post-treatment (Supplement 2, fig 1).

**Figure 2.**
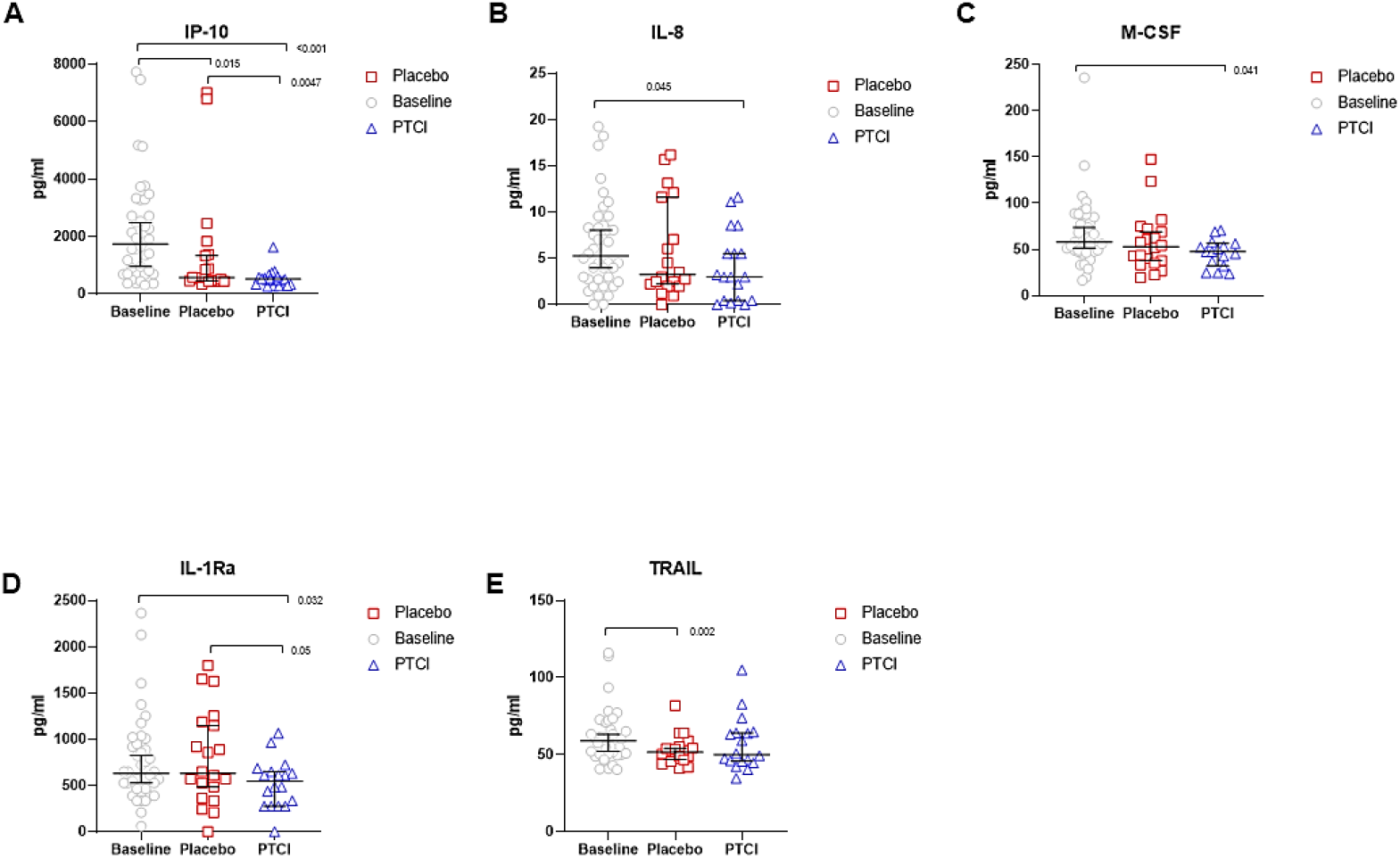
Serum cytokine and chemokine levels of SARS-CoV2-infected symptomatic outpatients at baseline and day 8 post-treatment with PTIC or placebo. Data are expressed as median with 95% confidence. Abbreviations: IP-10, IFN-γ inducible protein-10; IL-8, Interleukin-8; IL-1Ra, IL-1 receptor antagonist; M-CSF, Macrophage colony-stimulating factor; SARS-CoV-2, severe acute respiratory syndrome coronavirus 2; TRAIL, TNF-related apoptosis inducing ligand.

### Secondary outcomes

At days 8, 15 and 97 of follow-up, the percentage reported by the subjects with oxygen saturation readings ≥92% in the PTIC and placebo groups were 90 vs 67% (*P*=0.007), 98 vs 80% (*P*=0.009), and 100 vs 89% (*P*=0.033), respectively (table 2). The mean oxygen saturations in the PTIC and placebo groups in the aforementioned time points were 94±2.4 vs 93±3.3 (*P*=0.085), 95±1.7 vs 93±2.2 (*P*=0.003), and 95±2.1 vs 95±2.3 (*P*=0.429), respectively (Table 2).

**Table 2.** Study endpoints.

| Characteristic | 8-day post-treatment with |  |  | 15-days post-treatment with |  |  | 97-days post-treatment with |  |  |
| --- | --- | --- | --- | --- | --- | --- | --- | --- | --- |
|  | PTIC<br>(N=44) | Placebo<br>(N=43) | P-value | PTIC<br>(N=42) | Placebo<br>(N=39) | P-value | PTIC<br>(N=40) | Placebo<br>(N=37) | P-value |
| SpO2 ≥ 92%, n (%) | 40 (90.1) | 29 (67.4) | **0.007** | 41 (97.6) | 31 (79.5) | **0.009** | 40 (100) | 33 (89.2) | **0.033** |
| pSO<sub>2</sub>; mean ± SD | 94 ± 2.4 | 93 ± 3.3 | 0.085 | 95 ± 1.7 | 93 ± 2.2 | **0.003** | 95 ± 2.1 | 95 ± 2.3 | 0.429 |
| Median | 94 | 93 |  | 95 | 93 |  | 95 | 95 |  |
| IQR | 92-95 | 91-95 |  | 93-96 | 92-95 |  | 93-97 | 93-97 |  |
| O2 supplementation, n (%) | 2 (4.5) | 4 (9.3) | 0.381 | 1 (2.3) | 1 (2.6) | 0.958 | 0 (0.0) | 0 (0.0) | - |
| **Inpatient Admissions** | 0 (0.0) | 3 (7.0) | **0.075** | 0 (0.0) | 0 (0.0) | - | 0 (0.0) | 0 (0.0) | - |
| **Symptoms** |  |  |  |  |  |  |  |  |  |
| Dyspnea, n (%) | 6 (13.6) | 10 (25.6) | 0.166 | 3 (7.1) | 9 (23.1) | **0.044** | 6 (15) | 6 (16.2) | 0.883 |
| Δ (%) | -66.6 | -33.3 |  | -83.3 | -40 |  | -66.6 | -60 |  |
| Cough, n (%) | 17 (38.6) | 22 (56.4) | 0.105 | 11 (26.2) | 21 (53.8) | **0.011** | 4 (10) | 6 (16.2) | 0.418 |
| Δ (%) | -50 | -33.3 |  | -67.6 | -36.3 |  | -88.2 | -81.8 |  |
| Chest pain, n (%) | 8 (18.2) | 9 (23.1) | 0.581 | 5 (11.9) | 6 (15.4) | 0.648 | 7 (17.5) | 1 (2.7) | 0.033 |
| Δ (%) | -57.8 | -43.7 |  | -73.6 | -62.5 |  | -63.1 | -93.7 |  |
| Rhinorrhea, n (%) | 9 (20.5) | 9 (41) | 0.772 | 6 (14.3) | 6 (15.4) | 0.889 | 5 (12.5) | 3 (8.1) | 0.528 |
| Δ (%) | -52.6 | -55.0 |  | -68.4 | 0.0 |  | -73.6 | -85.0 |  |
| Headache, n (%) | 12 (27.3) | 16 (41) | 0.186 | 9 (21.4) | 15 (38.5) | 0.093 | 10 (25) | 14 (37.8) | 0.224 |
| Δ (%) | -45.4 | -33.3 |  | -59.0 | -37.5 |  | -54.5 | -41.6 |  |
| Sore throat, n (%) | 9 (30.5) | 10 (25.6) | 0.575 | 5 (11.9) | 6 (15.4) | 0.648 | 6 (15) | 7 (18.9) | 0.646 |
| Δ (%) | -55.0 | -52.3 |  | -75.0 | -71.4 |  | -70.0 | -66.6 |  |
| Malaise, n (%) | 16 (36.4) | 18 (46.2) | 0.365 | 12 (28.6) | 11 (28.2) | 0.971 | 11 (27.5) | 8 (21.6) | 0.374 |
| Δ (%) | -40.7 | -33.3 |  | -55.5 | -59.2 |  | -59.2 | -70.3 |  |
| Arthralgia, n (%) | 8 (18.2) | 8 (20.5) | 0.788 | 6 (14.3) | 6 (15.4) | 0.889 | 7 (17.5) | 8 (21.6) | 0.648 |
| Δ (%) | -55.5 | -69.2 |  | -66.6 | -76.9 |  | -61.1 | -69.2 |  |
| Myalgia, n (%) | 12 (27.3) | 11 (28.2) | 0.925 | 5 (11.9) | 6 (15.4) | 0.648 | 7 (17.5) | 3 (8.1) | 0.221 |
| Δ (%) | -47.8 | -56.0 |  | -78.2 | -76.0 |  | -69.5 | -88.0 |  |
| Brain fog, n (%) | 7 (15.9) | 12 (30.8) | 0.108 | 6 (14.3) | 7 (17.9) | 0.654 | 9 (22.5) | 10 (27) | 0.645 |
| Δ (%) | -72.0 | -33.3 |  | -76.0 | -61.1 |  | -64.0 | -44.4 |  |
| Ageusia, n (%) | 18 (40.9) | 13 (33.3) | 0.476 | 11 (26.2) | 8 (20.5) | 0.547 | 5 (12.5) | 4 (10.8) | 0.818 |
| Δ (%) | -37.9 | -31.5 |  | -62.0 | -57.8 |  | -82.7 | -78.9 |  |
| Anosmia, n (%) | 23 (52.3) | 13 (33.3) | 0.082 | 16 (38.1) | 9 (23.1) | 0.144 | 6 (15) | 2 (5.4) | 0.168 |
| Δ (%) | 23.33 | 35.0 |  | 46.6 | 55 |  | 80.0 | 90.0 |  |
| Diarrhea, n (%) | 4 (9.1) | 6 (15.4) | 0.379 | 3 (7.1) | 2 (5.1) | 0.707 | 1 (2.5) | 0 (0.0) | 0.333 |
| Δ (%) | -63.63 | -25 |  | -72.7 | -75 |  | -90.9 | -100.0 |  |
| Abdominal pain, n (%) | 5 (11.4) | 6 (15.4) | 0.590 | 0 (0.0) | 3 (7.7) | 0.067 | 1 (2.5) | 3 (8.1) | 0.268 |
| Δ (%) | -37.5 | -57.1 |  | -100.0 | -78.5 |  | -87.5 | -78.5 |  |
| Jaundice, n (%) | 0 (0.0) | 2 (5.1) | 0.128 | 0 (0.0) | 0 (0.0) | - | 0 (0.0) | 1 (2.7) | 0.295 |
| Δ (%) | -100.0 | 100.0 |  | -100.0 | -100.0 |  | -100 | 0.0 |  |
| Vomiting and Nausea, n (%) | 0 (0.0) | 0 (0.0) |  | 1 (2.4) | 0 (0.0) | 0.332 | 0 (0.0) | 0 (0.0) | - |
| Δ (%) | -100.0 | -100.0 |  | -50 | -100.0 |  | -100.0 | -100.0 |  |
| Conjunctivitis, n (%) | 1 (2.3) | 1 (2.6) | 0.931 | 1 (2.4) | 1 (2.6) | 0.958 | 2 (5.0) | 1 (2.7) | 0.603 |
| Δ (%) | -88.88 | -90.9 |  | -88.8 | -90.9 |  | -77.7 | -90.9 |  |
| Cyanosis, n (%) | 0 (0.0) | 1 (2.6) | 0.285 | 0 (0.0) | 0 (0.0) | - | 0 (0.0) | 0 (0.0) | - |
| Δ (%) | 0.0 | 100.0 |  | 0.0 | 0.0 |  | 0.0 | 0.0 |  |

Δ: Delta calculated by taking: [(Baseline data - Day 1, 8, or 97 of follow-up) / Baseline data from table 1]\*100. IQR: interquartile range; PTCl: polymerized type I collagen; pSO<sub>2</sub>: oxygen saturation; SD: standard deviation *P* value: PTCl vs. Placebo

The Kaplan-Meier survival curve for oxygen saturations 92% or greater while breathing ambient air was statistically different between groups (log-rank *P* = 0.0109; fig 3A). Since there were no significant differences between groups at baseline, we did not make any adjustments. The Cox regression model indicated that the hazard for meeting an oxygen saturation lower than 92% was significantly lower in the PTIC than the placebo group (HR 0.25, Wald *P*-value=0.0384). When stratifying by age, no changes occurred. Based on the accelerated time failure model, subjects of the PTIC group reached oxygen saturations 92% or greater 2.7-fold faster than the placebo group at 3 and 8 days (*P*<0.001 in both cases). In terms of risk, this implied that the PTIC group had a 63% lower risk for mean oxygen saturations readings below 92% (*P* <0.001; fig 3B).

**Figure 3.**
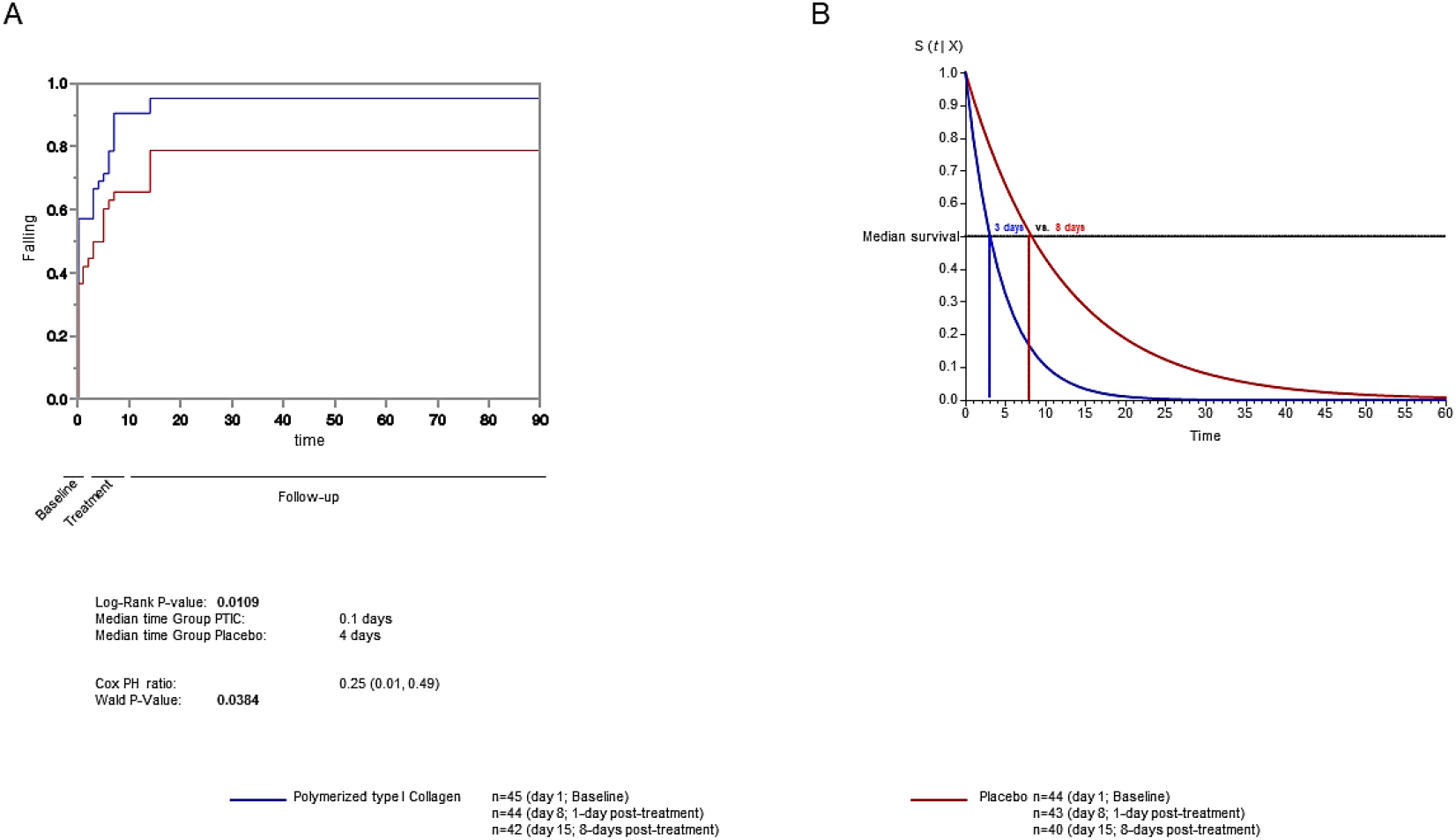
(A) Probability of oxygen saturation 92% or greater while breathing ambient air. (B) Accelerated time failure model for oxygen saturation 92% or greater while breathing ambient air among polymerized type I collagen and placebo.

Symptom improvement was reported daily by every patient and compared with baseline (fig 4). At days 8 and 15 post-treatment with PTIC, 66.6% and 83% of subjects no longer had dyspnoea vs baseline, respectively (*P*=0.044; table 2); corresponding values in the placebo group were 33.3% and 40%, respectively (table 2). Compared with baseline, significant improvements in dyspnoea intensity were noticed at day 4 of follow-up in PTIC subjects, as compared to day 97 for placebo subjects (fig 4A).

**Figure 4.**
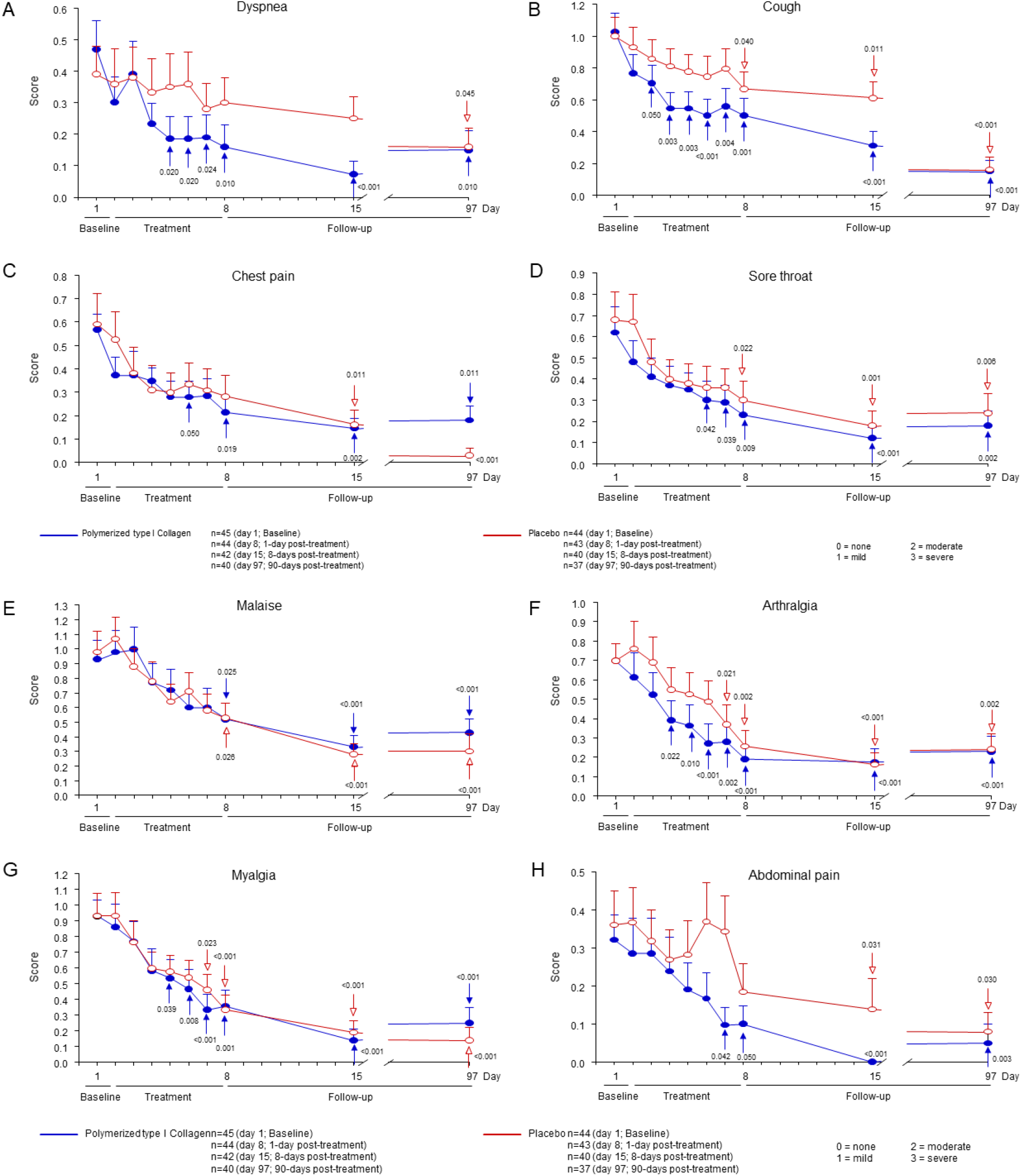
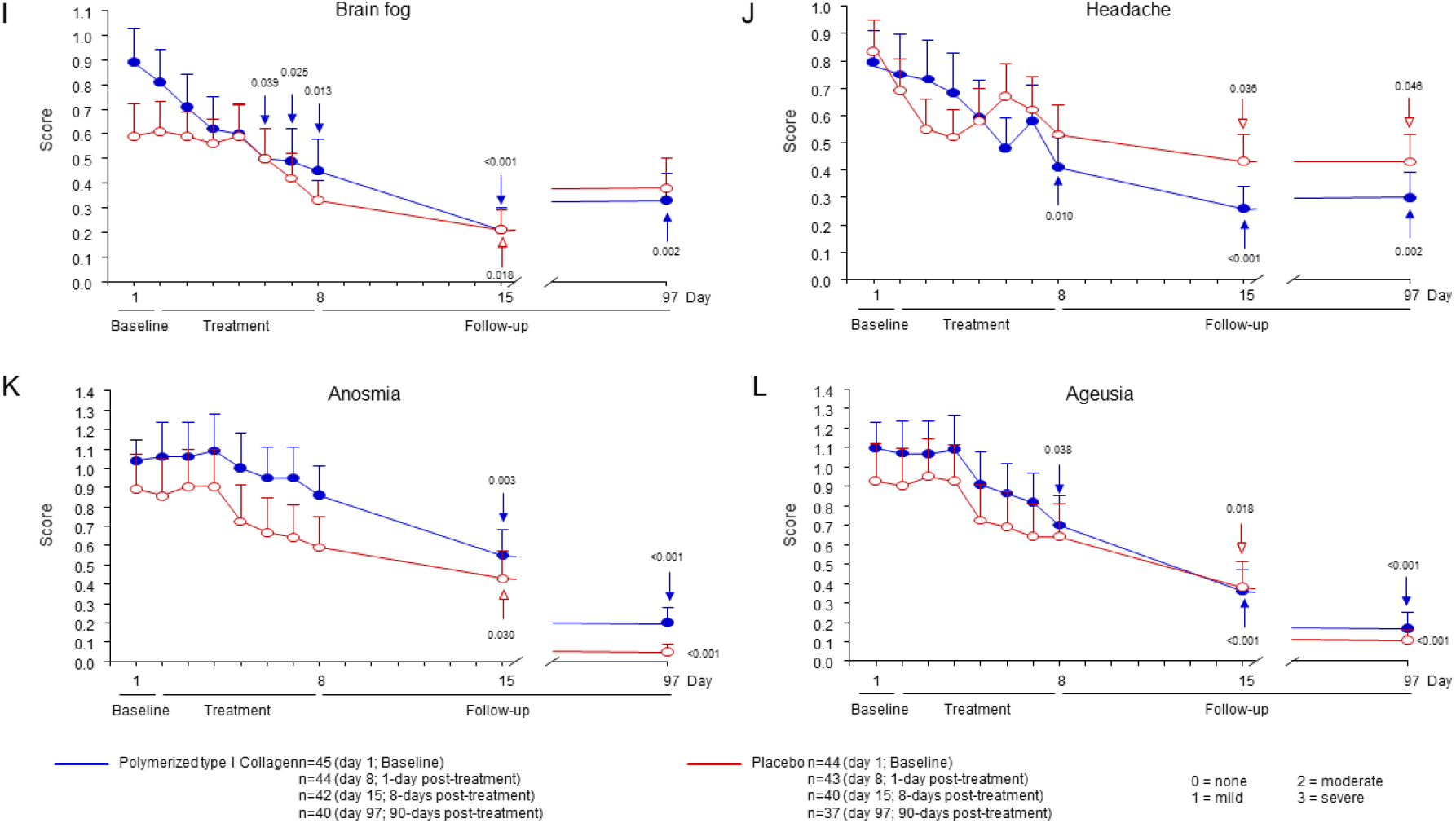
Intensity of symptoms during treatment and follow-up of outpatients with symptomatic COVID-19 treated with polymerized type I collagen or placebo. (A) Dyspnoea, (B) cough, (C) chest pain, (D) sore throat, (E) malaise, (F) arthralgia, (G) myalgia, (H) abdominal pain, (I) brain fog, (J) headache, (K) anosmia, and (L) ageusia. The intensity of the symptom was evaluated on a 4-point rating scale (0 = without symptom, 1 = mild, 2 = moderate, 3 = severe). Blue lines represent the group of patients under polymerized type I collagen treatment. Red lines represent the group of patients under placebo treatment. Results depict mean ± standard error of the mean. Blue arrows show the day in which the treatment reached a *P* < 0.05 compared to baseline for polymerized type I collagen treatment. Red arrows show the day in which the treatment reached a *P* < 0.05 compared to baseline for placebo.

At days 8 and 15 post-treatment with PTIC, 50.0% and 67.6% of the patients no longer had cough vs baseline, respectively (*P*=0.011; table 2); corresponding values in the placebo group were 33.3% and 33.6%, respectively (table 2). Cough intensity decreased significantly after 2 days of treatment with PTIC as compared to 8 days with placebo (fig 4B).

Arthralgia and myalgia intensity decreased significantly after 4-5 days of treatment with PTIC vs 7 days with placebo (fig 4F and G).

Chest pain, sore throat and brain fog diminished after 6 days of treatment with PTIC vs 8-15 days with placebo (Fig 4C). Abdominal pain, headache and ageusia diminished after 7-8 days of treatment with PTIC as compared to 15 days with placebo (fig 4H, J and L).

Nonetheless, subjects in both the PTIC and placebo groups reported at least one long-lasting residual symptom during follow-up, including dyspnoea (15% vs 16%), brain fog (22.5 vs 27%), headache (25 vs 37.8%), pain throat (15 vs 18.9%), malaise (27.5% vs 21.6%), chest pain (17.5 vs 2.7%), arthralgias (17.5% vs 21.6%) and myalgias (17.5 vs 8.1%), respectively (table 2, fig 4).

### Invasive mechanical ventilation, supplemental oxygen, hospitalizations and deaths

At day 8 post-treatment, 6 of 87 patients (7%) received supplemental oxygen via nasal cannula without the need for invasive mechanical ventilation at home: 2/44 (4.5%) of the PTIC group (one patient received 2L/min and another one received 3L/min), and 4/43 (9.3%) of the placebo group (4-10L/min) (table 2).

At day 15 post-treatment, 2 of 81 patients (2.5%) received supplemental oxygen via nasal cannula without the need for invasive mechanical ventilation: 1/42 (2.3%) of the PTIC group (one patient received 2L/min), and 1/39 (2.6) of the placebo group (4L/min) (table 2).

At day 97 post-treatment, none of the patients required supplemental oxygen (table 2).

At 8-day post-treatment, 3 of 43 subjects (7%) of the placebo group were hospitalized for 5-21 days (Table 2). All patients were discharged alive and no deaths occurred. No subjects in the PTIC treatment group were hospitalized.

### Laboratory assays

No differences in laboratory results were found among the PTIC and placebo groups at baseline (Table 1).

At day 8 and 15 post-treatment with PTIC, serum levels of high sensitivity CRP (hsCRP) decreased (52% and 73%, respectively) compared with baseline levels (*P*=0.002 and *P*<0.001). In the placebo group, hsCRP levels were 3% and 67% lower at 1 and 8 days compared with baseline levels (Supplement 2, fig 2A; table 3).

**Table 3.**
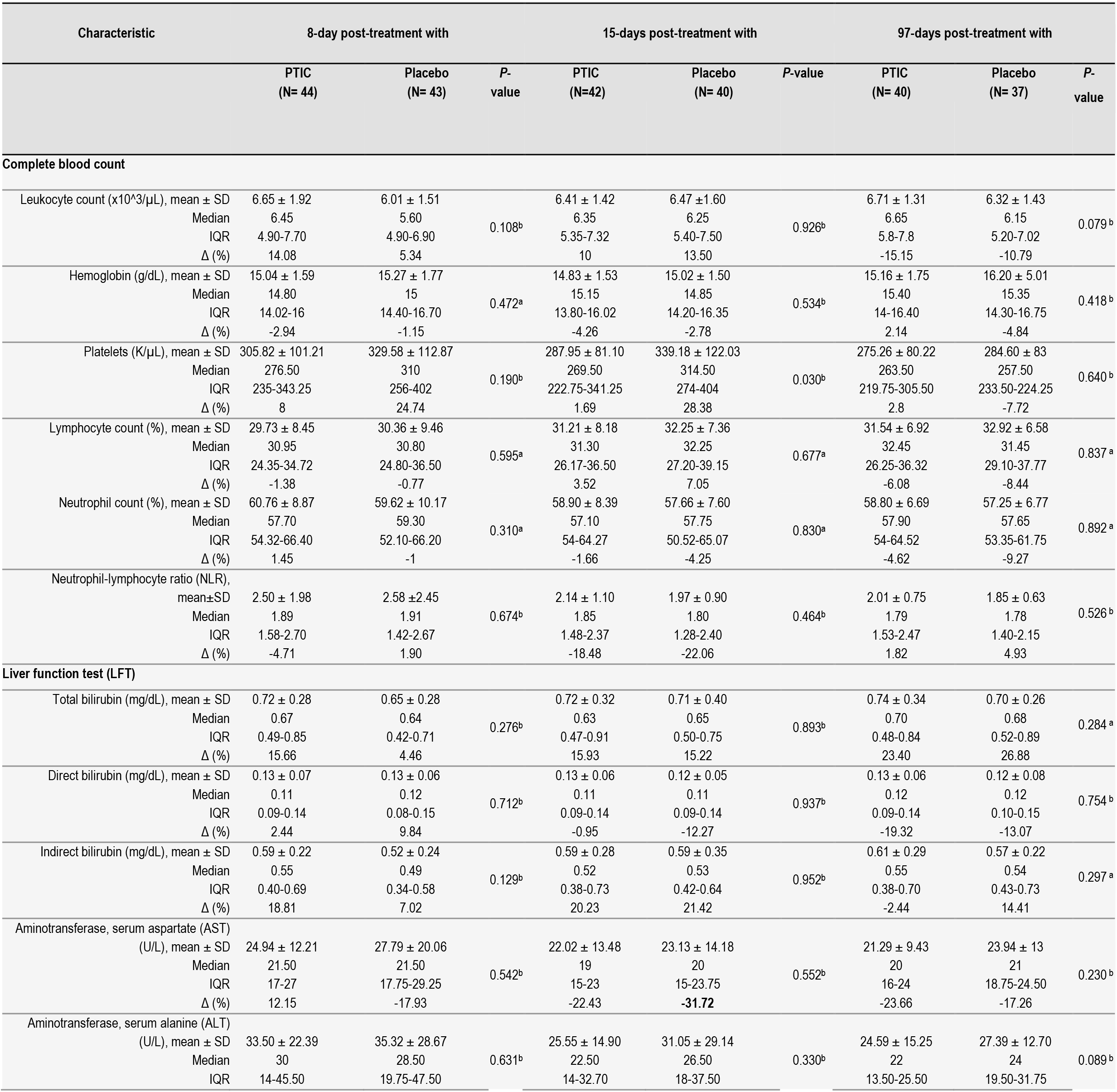
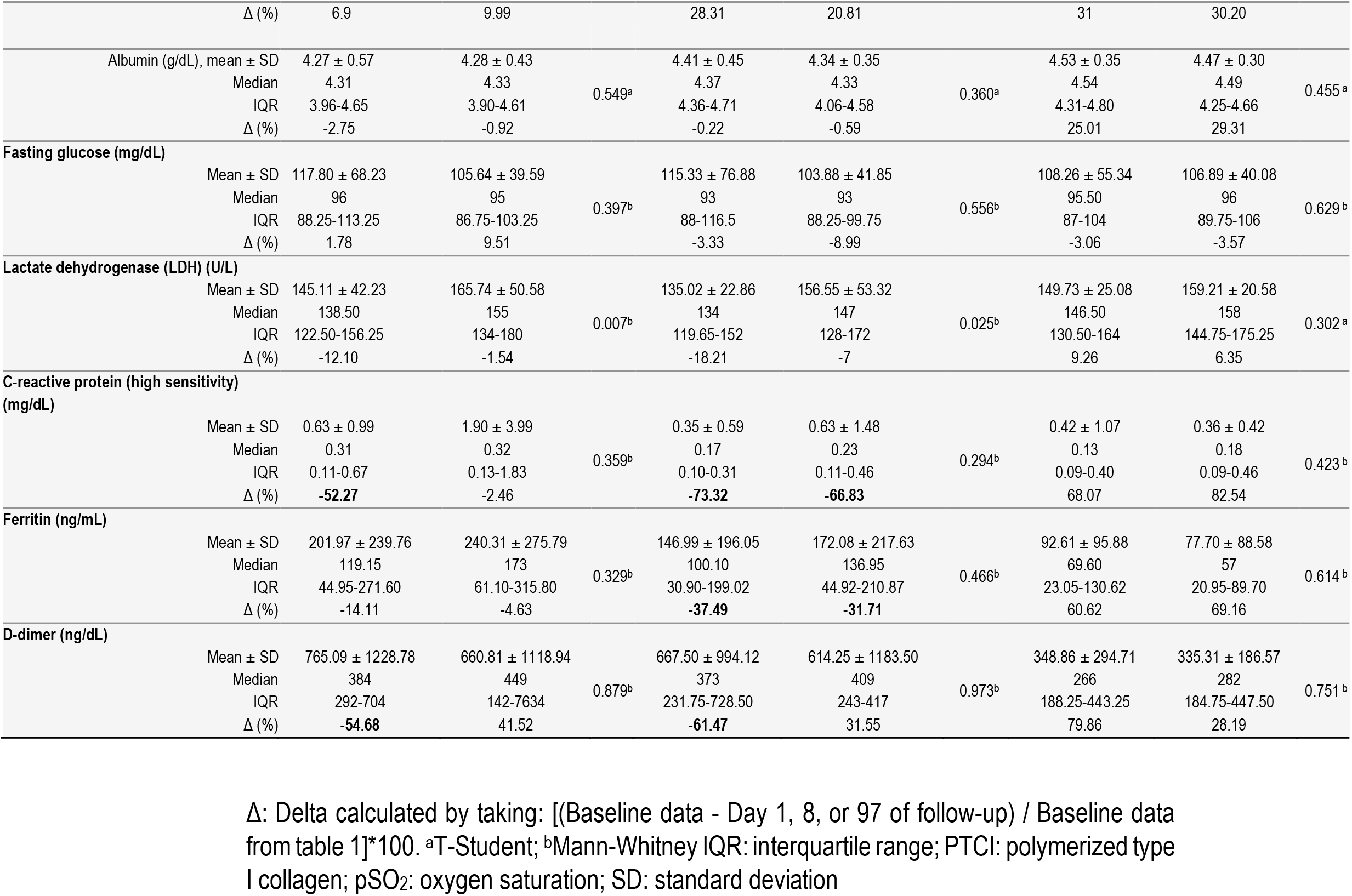
Follow-up of laboratory tests.

At day 8 and 15 post-treatment, D-dimer levels in PTIC subjects decreased (55% and 61%, respectively); in the placebo group, D-dimer increased 42% and 32%, respectively (Supplement 2, fig 2B; table 3). No differences were detected in the other laboratory variables compared to the baseline.

### Chest imaging

Imaging of subjects initially revealed characteristic patchy infiltration, progressing to large ground-glass opacities that often presented bilaterally. Abnormalities on chest CT scans were detected among 82% of the patients included in this study, and no differences between groups were detected (table 1).

### Adverse events

In this study, no serious adverse events were detected. PTIC was safe and well tolerated. In the PTIC group, the following were observed at day 1 after treatment: 31 patients had pain in the injection site lasting 15-20 minutes, 1 patient complained of drowsiness, and 1 patient had an urticarial rash at the injection site on day 6. At day 8, 1 patient had a right hemicrania headache with photophobia that improved with acetaminophen.

In the placebo group, the following were observed at day 1 after treatment: 28 patients had pain in the injection site lasting 15-20 minutes; 1 patient had facial rash (day 2 of treatment), muscle contracture in the lumbar area (day 3 of treatment), and anterior chest paraesthesia (day 4 of treatment); 1 patient had abdominal pain; and 1 patient had chest pain and vomiting. At day 8, 1 patient had nocturnal headache with parasympathetic symptoms that woke her up for 15 to 20 min, 1 patient had tachycardia, and 1 patient had chest pain and oral candidiasis.

No adverse events were detected at day 97 of follow-up.

## Discussion

Clinical and physiologic studies of COVID-19 have suggested a biphasic pattern of illness with an initial viral response phase lasting 7 to 10 days after symptom onset characterized by rapid viral replication followed by an inflammatory response phase driven by innate and adaptive immune responses to the virus. In a subset of individuals, immune dysregulation leads to a catastrophic maladaptive hyperimmune response that is strongly linked to increased morbidity and mortality. CRS can develop a hemophagocytic lymphohistiocytosis or macrophage activation syndrome-like syndrome with the rise of other cytokines and chemokines (between 6 to 8 days after onset of the disease) including IL-1β, IL-1 receptor antagonist (IL-1RA), IL-7, IL-8, IL-9, IL-10, G-CSF, GM-CSF, IFN-γ, IP-10, MCP-1, MIP-1α, and MIP-1β.^3,21,23,27^ In addition, it has been shown that IP-10, MCP-3, HGF, MIG, IL-1Ra, and MIP-1α expression levels are excellent biomarkers for the prediction of COVID-19 progression and severity.^3,21,27-30^ They highly and positively correlate with D-dimer and Murray scores, with *ρ* values greater than 0.45 and *P* values less than 0.0001, whereas IL-1Ra, IL-2Ra, IL-6, IL-10, IL-18, M-CSF, and IFN-γ show weaker, albeit significant, positive associations with Murray scores.^21^ Moreover, it has been demonstrated that the continuously high level of these cytokines and chemokines are associated with deteriorated progression of disease and fatal outcome.

IL-8 can contribute to the destabilization of endothelial cells in CRS, leading to the development of vascular barrier impairment and capillary damage with multiorgan failure.^26^ IL-8 levels are higher in COVID-19 patients with mild symptoms than in healthy people and further elevated in severe COVID-19 patients, for this reason may serve as a biomarker to indicate the COVID-19 disease prognosis.^31^

Avoiding that patients progress to CRS has important therapeutic implications, since anti-inflammatory agents are likely to be more useful than anti-virals in this setting.^26^ In this study, it has been demonstrated that PTIC treatment was useful for decreasing the levels of IP-10, IL-8, and M-CSF, all of them biomarkers of severe disease, during de first week of treatment. This finding is relevant, since Zhao Y., et al., have described that in the group of patients with mild disease, IP-10 levels decline from week 2 and return to normal in week 4, while in the severe cases the IP-10 levels remain high level at week 2 and start to decline at week 3 and further by week 4.^32^ Therefore, treatment with PTIC reduces the levels of IP-10 by 70% at week 1 in patients with moderate disease, suggesting its regulatory role on CRS. Finally, TRAIL has been involved in the innate immune response to infection and has been shown to correlate with disease severity,^30^ thus patients admitted to ICU have lower average TRAIL level compared to patients not admitted to ICU. In our cohort, patients treated with placebo had lower TRAIL levels vs baseline.

Intramuscular PTIC was associated with better oxygen saturation values when compared to placebo. Also, PTIC shortened symptom duration. At day 8 and 15 post-treatment with PTIC, a higher mean oxygen saturation value and a higher proportion of patients retaining oxygen saturation values ≥92% were observed. This could be related to a decrease in dyspnoea and chest pain, as well as cough. However, at least one residual symptom was reported at follow-up, as previously published.^33^

Regarding systemic inflammation, at day 8 and 15 post-treatment with PTIC, statistically significant lower levels of hsCRP and D-dimer were observed. The benefit was clear in the early stage of the infection (7 days after symptom onset). A 3- to 4-fold increase in levels of D-dimer in the early stages of COVID-19 disease is linked to poor prognosis.^34^ Moreover, CRP reflects total systemic burden of inflammation in several disorders. CRP has been shown to upregulate the production of proinflammatory cytokines and adhesion molecules (ICAM-1, VCAM-1 end ELAM-1), and its expression is regulated by a proinflammatory milieu enriched with IL-6.^35^ High levels of CRP are found closely correlated with disease severity.^38^ Based on the reduction of hsCRP and D-dimer levels in patients under treatment with PTIC, we consider that it has anti-inflammatory properties, as seen with dexamethasone, colchicine, and Janus kinase inhibitors.^4.36-41^ Nonetheless, in contrast to dexamethasone, PTIC does not affect the functions of T and B cells, as previously reported;^16^ in consequence, PTIC would not be expected to affect serum viral load or the risk of acquiring other infectious agents, nor to lead to water and salt retention, blood pressure rises, hyperglycaemia, muscle weakness, gastrointestinal bleeding or psychological disturbances, as observed with corticosteroids,^41^ although this must be confirmed in further studies. Moreover, PTIC was safe, well-tolerated and effective for improving symptoms in outpatients with mild to moderate COVID-19. It did not induce liver damage, impairment of haematopoiesis or alterations in blood count.

Most COVID-19 patients show a benign disease and overcome viral inflammation by robust but not overreactive immune responses.^42^ Thus, few studies have focused on this vulnerable population.

We think that treating outpatients with PTIC could potentially avoid visits to the Emergency Department and hospitalizations, and as judged by symptom improvement, could aid in prevention of sequelae, such as persistent dyspnoea.

In the present study, 7% of patients that received placebo were hospitalized and received non-invasive high flow oxygen through a nasal cannula (10L/min) and dexamethasone. They were discharged alive.

The potential advantages of PTIC for the treatment of outpatients with COVID-19 are its safety profile, low cost, and intramuscular administration, as compared to REGN-COV2 (neutralizing monoclonal antibody cocktail), which is administered intravenously to mild/moderate COVID-19 patients.^43^

We acknowledge several limitations. This is a small study conducted within a single centre, so findings should be regarded as preliminary until replicated in larger clinical trials with a more heterogeneous study population. Clinical outcomes were only exploratory. The self-reporting symptom severity, or rather, the lack of information to verify compliance, is one of the study’
ss limitation. The possible usefulness of PTIC for treatment of hospitalized and/or patients with invasive ventilation is still unknown. We did not measure viral loads, so a possible effect of PTIC on viral kinetics cannot be ruled out. Finally, the underlying mechanisms for the beneficial effects of PTIC observed in this study need further clarification.

In summary, in this clinical trial of adult outpatients with mild to moderate symptomatic COVID-19, intramuscular PTIC was safe and well tolerated, downregulated CRS, efficiently shortened symptom duration, and was associated with better oxygen saturation values when compared to placebo.

## Data Availability

Data Sharing Statement: In accordance with NHS Digital Information Governance requirements, the study data cannot be shared. The manuscript guarantor affirms that this manuscript is an honest, accurate, and transparent account of the study being reported; that no important aspects of the study have been omitted; and that any discrepancies from the study as planned (and, if relevant, registered) have been explained.

## Author Contributions

Furuzawa-Carballeda and Torres-Villalobos had full access to all of the data in the study and take responsibility for the integrity of the data in the accuracy of the data analysis.

### Concept and design

Furuzawa-Carballeda and Torres-Villalobos

### Acquisition, analysis, or interpretation of data

Méndez-Flores, Priego-Ranero, Azamar-Llamas, Olvera-Prado, Rivas-Redondo, Ochoa-Hein, Rojas-Castañeda, Urbina-Terán, Septién-Stute, Hernández-Gilsoul, Aguilar-Morgan, Olivares-Martínez, and Hernández-Ramírez.

### Drafting of the manuscript

Furuzawa-Carballeda, Ochoa-Hein and Torres-Villalobos.

### Critical revision of the manuscript for important intellectual content

Méndez-Flores, Priego-Ranero, Azamar-Llamas, Olvera-Prado, Rivas-Redondo, Ochoa-Hein, Rojas-Castañeda, Olivares-Martínez, Hernández-Ramírez, Furuzawa-Carballeda and Torres-Villalobos.

### Statistical analysis

Fernández-Camargo, and Perez-Ortiz.

### Obtained funding

Furuzawa-Carballeda

### Supervision

Furuzawa-Carballeda, and Torres-Villalobos

## Ethics declarations

The study was approved by the institutional review board at Instituto Nacional de Ciencias Médicas y Nutrición Salvador Zubirán (INCMNSZ, reference no. IRE 3412-20-21-1) and was conducted in compliance with the Declaration of Helsinki (World Medical Association. World Medical Association Declaration of Helsinki. *JAMA*. 2013;310(20):2191-2194.), the Good Clinical Practice guidelines, and local regulatory requirements. All participants provided written informed consent.

## Role of the founding source

The funder of the study had no role in the study design, data collection, data analysis, data interpretation, or writing of the report. The corresponding authors had full access to all the data in the study and had final responsibility for the decision to submit for publication.

## Data Sharing Statement

In accordance with NHS Digital’s Information Governance requirements, the study data cannot be shared. The manuscript’s guarantor affirms that this manuscript is an honest, accurate, and transparent account of the study being reported; that no important aspects of the study have been omitted; and that any discrepancies from the study as planned (and, if relevant, registered) have been explained.

## Additional Contributions

Polymerized type I collagen was donated by Aspid SA de CV. We thank Dr. Alicia Frenk-Mora, Dr. Judith González-Sánchez, and MSc. Ivonne Aidé Lomelí Almanza, for their valuable assistance with the organization of patient appointments. We thank all patients involved in the study. We also thank the Triage and Emergency Departments.

BMI: body mass index; IQR: interquartile range; PTCI: polymerized type I collagen; pSO_2_: oxygen saturation; SD: standard deviation

