## Supplementary material for "Effect of polymerized type I collagen in hyperinflammation of adult outpatients with symptomatic COVID-19: a double blind, randomised, placebo-controlled clinical trial": Suplemental figures

**eFigure1.** Serum cytokine and chemokine levels of SARS-CoV2-infected symptomatic outpatients at baseline and day 1 post-treatment with PTIC or placebo. Data are expressed as median with 95% confidence

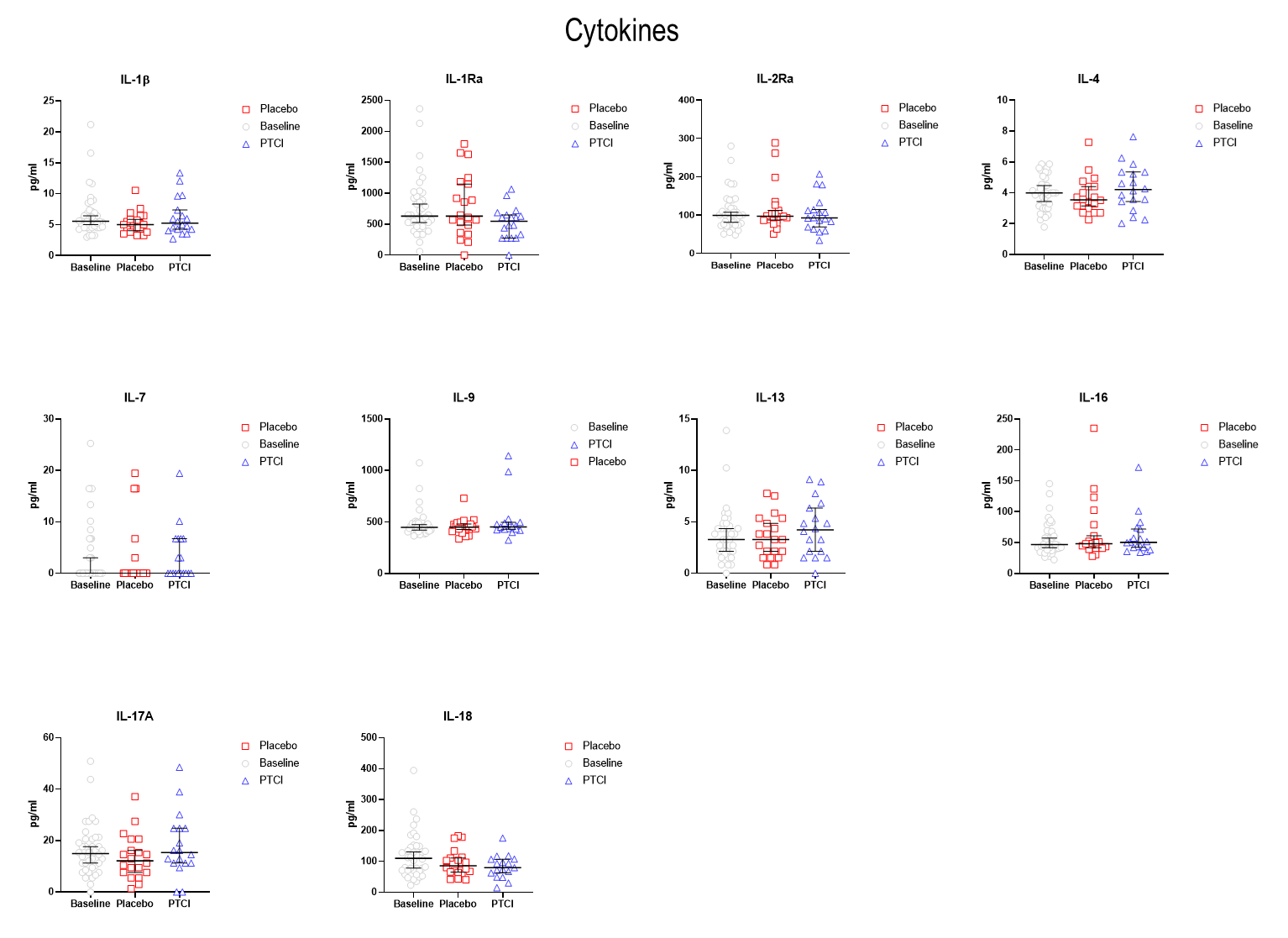

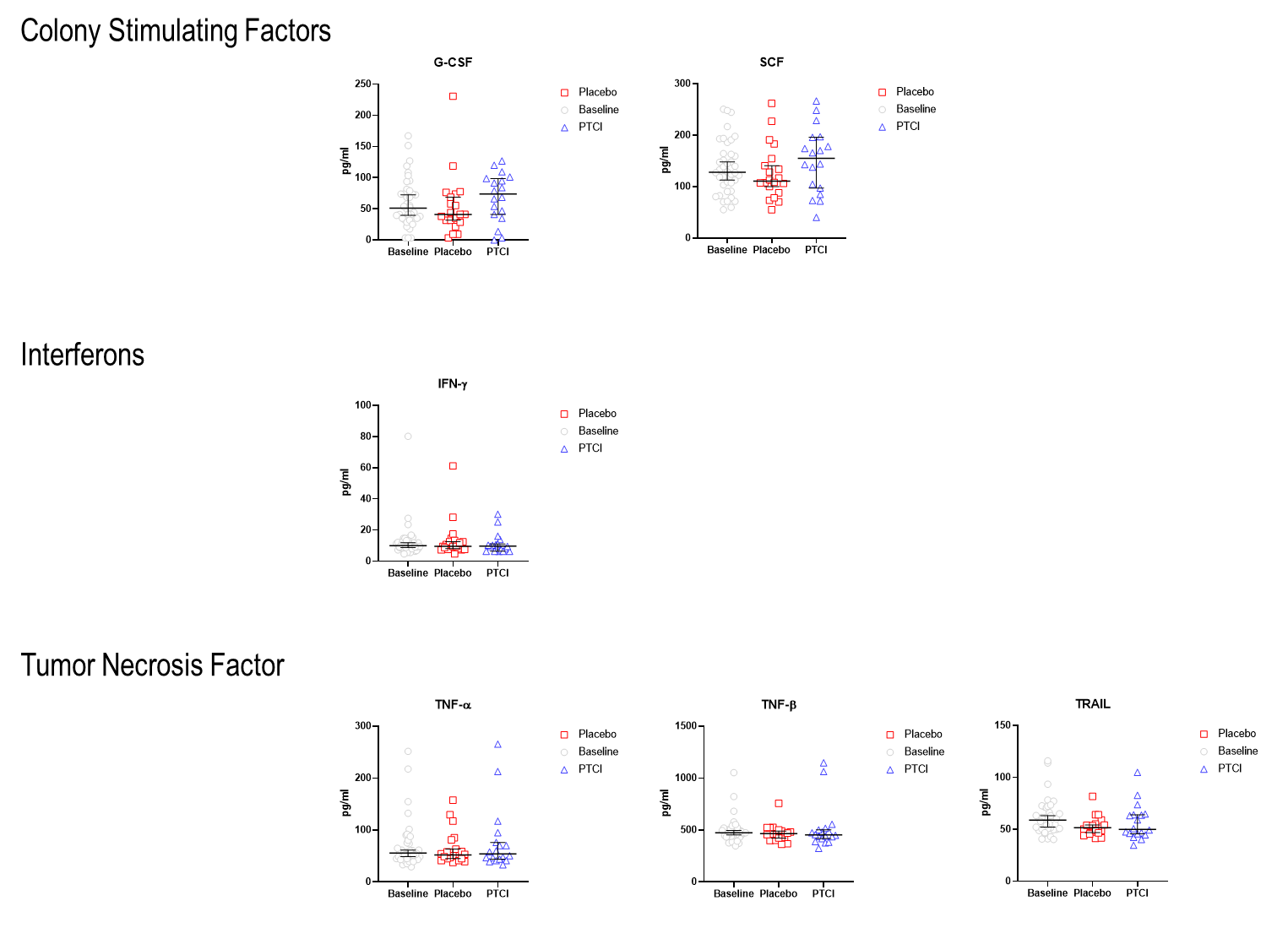

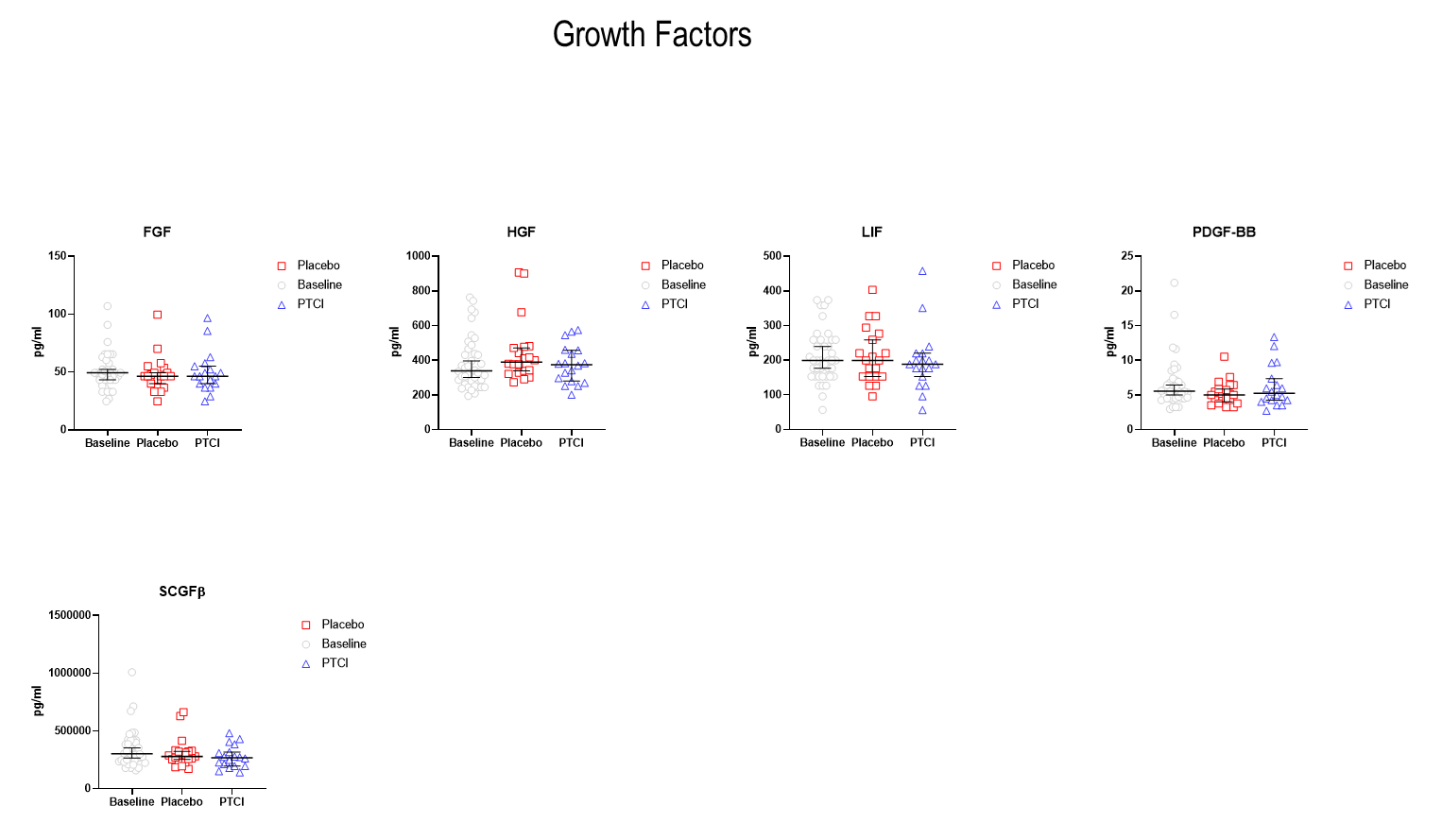

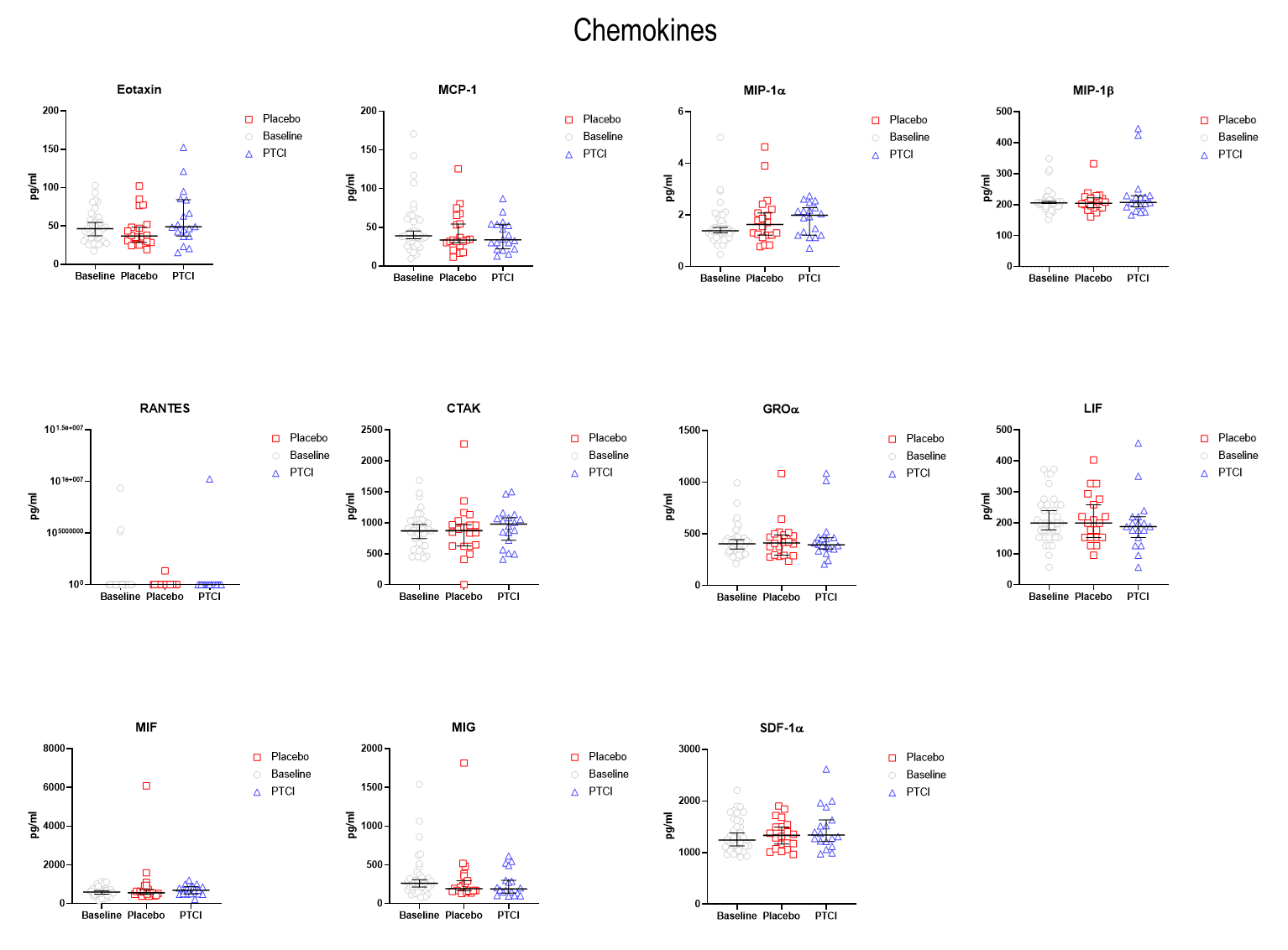

**eFigure 2.** PCR and D-dimer during treatment and follow-up of outpatients with symptomatic COVID-19 treated with polymerized type I collagen or placebo. (A) high sensitivity C-reactive protein, and (B) D-dimer. Blue lines represent the group of patients under polymerized type I collagen treatment. Red lines represent the group of patients under placebo treatment. Results depict mean ± error standard of the mean. Blue and red arrows show *P-value* (Day 1, 8 and 97 days post-treatment compared to baseline).

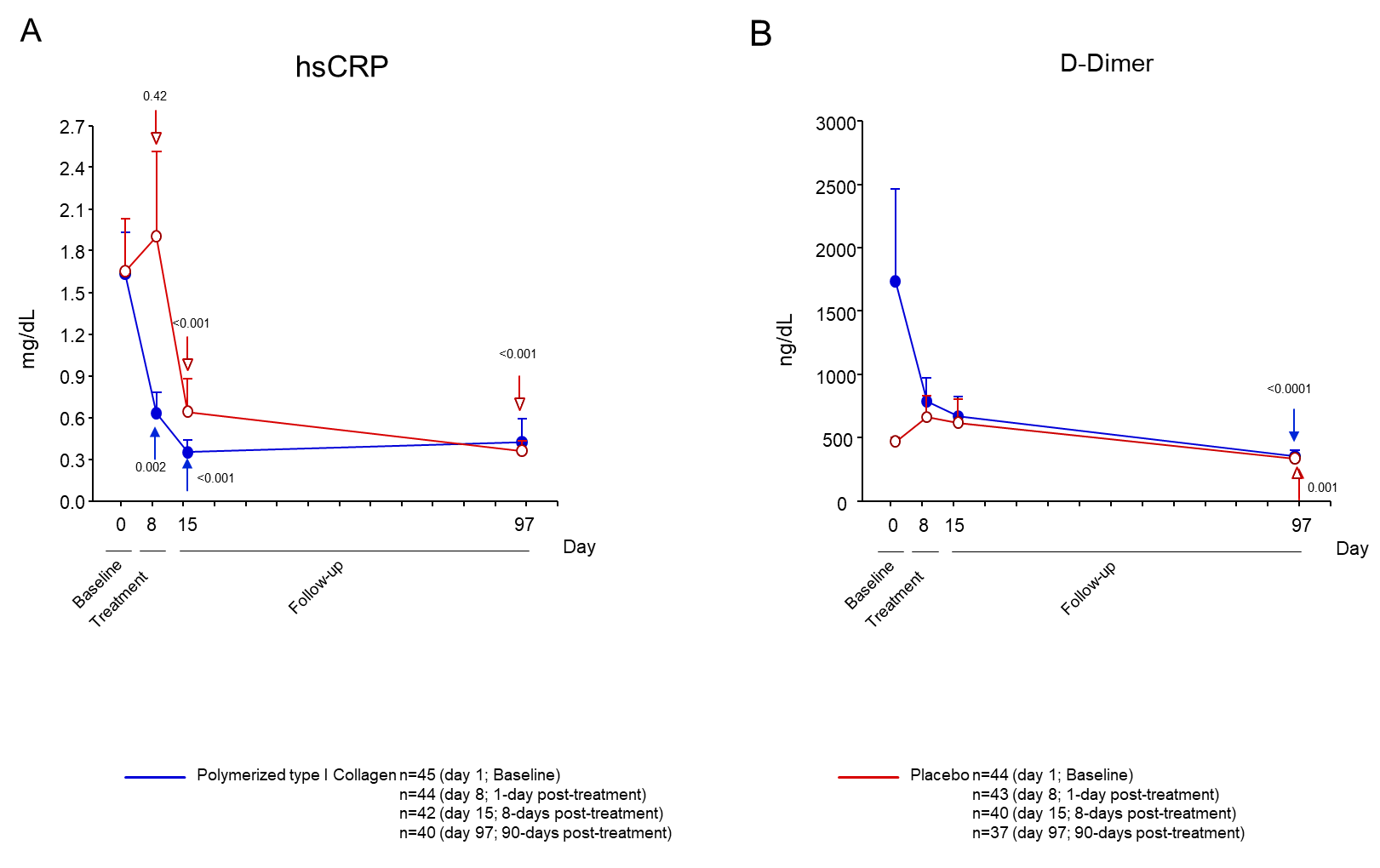

| eTable 1. Non-serious adverse events | |
| --- | --- |
| **MedDRA System Organ Class** | **Preferred Term** |
| Any System Organ Class | Any Preferred Term |
| Blood and lymphatic system disorders | Anemia |
|  | Lymphopenia |
| Cardiac disorders | Atrial fibrillation |
| General disorders and administration site conditions | Pyrexia |
| Infections and infestations | Pneumonia |
| Investigations | Hemoglobin decreased |
|  | Glomerular filtration rate decreased |
|  | Aspartate aminotransferase increased |
|  | Lymphocyte count decreased |
|  | Blood glucose increased |
|  | Alanine aminotransferase increased |
|  | Blood bilirubin increased |
|  | Blood creatinine increased |
|  | Prothrombin time prolonged |
|  | Blood albumin decreased |
|  | Transaminases increased |
|  | Creatinine renal clearance decreased |
| Metabolism and nutrition disorders | Hyperglycemia |
|  | Acidosis |
|  | Hypoalbuminemia |
|  | Alkalosis |
| Psychiatric disorders | Delirium |
| Renal and urinary disorders | Acute kidney injury |
| Respiratory, thoracic and mediastinal disorders | Hypoxia |
|  | Dyspnea |
|  | Respiratory distress |
| Uncoded | Uncoded |
| Vascular disorders | Hypotension |
|  | Hypertension |
|  | Deep vein thrombosis |

| eTable 2. Serious adverse events | |
| --- | --- |
| **MedDRA System Organ Class** | **Preferred Term** |
| Any System Organ Class | Any Preferred Term |
| Cardiac disorders | Cardiac arrest |
|  | Atrial fibrillation |
| Infections and infestations | Septic shock |
|  | Pneumonia viral or bacterial |
| Investigations | Glomerular filtration rate decreased |
| Renal and urinary disorders | Acute kidney injury |
| Respiratory, thoracic and mediastinal disorders | Respiratory failure |
|  | Acute respiratory failure |
|  | Respiratory distress |
|  | Hypoxia |
|  | Pneumothorax |
|  | Pulmonary embolism |
| Surgical and medical procedures | Mechanical ventilation |
|  | Endotracheal intubation |
| Uncoded | Uncoded |
| Vascular disorders | Hypotension |
|  | Shock |
